# Accuracy of sensor-based classification of clinically relevant motor activities in daily life of children with mobility impairments

**DOI:** 10.1101/2022.08.01.22278307

**Authors:** Fabian Marcel Rast, Florence Jucker, Rob Labruyère

## Abstract

**Background:** Wearable inertial sensors enable objective, long-term monitoring of motor activities in the children’s habitual environment after rehabilitation. However, sophisticated algorithms are needed to derive clinically relevant outcome measures. Therefore, we developed three independent algorithms based on the needs of pediatric rehabilitation. The first algorithm estimates the duration of lying, sitting, and standing positions and the number of sit-to-stand transitions with data of a trunk and a thigh sensor. The second algorithm detects active wheeling periods and distinguishes it from passive wheeling with data of a wrist and a wheelchair sensor. The third algorithm detects walking periods, discriminates between free and assisted walking, and estimates the covered altitude change during stair climbing with data of a single ankle sensor and a sensor placed on walking aids.

**Research question:** This study aimed to determine the accuracy of each algorithm in children undergoing rehabilitation.

**Methods:** Thirty-one children and adolescents with various medical diagnoses and levels of mobility impairments performed a semi-structured activity circuit. They wore inertial sensors on both wrists, the sternum, and the thigh and shank of the less-affected side. Video recordings, which were labeled by two independent researchers, served as reference criteria to determine the algorithms’ performance.

**Results:** The activity classification accuracy was 97% for the posture detection algorithm, 96% for the wheeling detection algorithm, and 93% for the walking detection algorithm.

**Significance:** This study presents three novel algorithms that provide a comprehensive and clinically relevant view of the children’s motor activities. These algorithms are described reproducibly and can be applied to other inertial sensor technologies. Moreover, they were validated in children with mobility impairments and can be used in clinical practice and clinical trials to determine the children’s motor performance in their habitual environment. To enable the evaluation of future algorithms, we published the labeled dataset.

## Background

Pediatric rehabilitation aims to foster functional independence in everyday life activities of children with congenital and acquired illnesses and injuries. For most families with children undergoing rehabilitation, improvements in self-care and mobility activities are prioritized [1]. Thereby, most rehabilitation goals are set about changing and maintaining body positions or walking and moving [2]. Hence, assessments covering these domains are essential to tailor therapy to the families’ needs and monitor the children’s progress over time.

In clinical practice, assessments are usually conducted in a standardized environment. Therefore, their outcomes reflect the children’s highest probable level of functioning within this setting (i.e., motor capacity) [3]. However, it has been shown that capacity only partially explains how the children perform in their habitual environment after rehabilitation (i.e., motor performance) [4,5]. Consequently, there is a need to measure performance directly by bringing assessments into daily life.

Recent advances in wearable sensor technologies overcome the limitation mentioned above by enabling objective and long-term monitoring of motor activities in a patient’s habitual environment [6]. Recent studies validated such sensor technologies and their underlying data processing algorithms in children with mobility impairments [7–10]. However, these algorithms do not provide a comprehensive view of the patients’ activities, and they lack an estimation of clinically relevant outcome measures. Moreover, they were only validated in children with cerebral palsy and spina bifida, representing approximately one third of all children undergoing rehabilitation [2]. Hence, the validity in children with other mobility impairments is still unknown.

Here, we present a novel algorithm that was developed based on the findings of two preceding studies assessing the clinical needs of pediatric rehabilitation. The first study was an international survey with pediatric healthcare professionals, and the second study investigated the frequency of rehabilitation goals at our center [2]. The results revealed the demand to have three sub-algorithms that require different sensor-setups and can be used independently: The first sub-algorithm estimates the duration of lying, sitting, and standing positions and the number of sit-to-stand transitions with data of a trunk and a thigh sensor. The second sub-algorithm detects active wheeling periods and distinguishes them from passive wheeling with data of a wrist and a wheelchair sensor. The third sub-algorithm detects walking periods, discriminates between free and assisted walking, and estimates the covered altitude change during stair climbing with data of a single ankle sensor and a sensor placed on walking aids. The aim of this study was to validate the three sub-algorithms in children undergoing rehabilitation. Specifically, we investigated the algorithm’s activity classification accuracy and determined the measurement error of the outcome measures.

## Method

### Participants & recruitment

A convenience sample of 31 children and adolescents was recruited at the Swiss Children’s Rehab of the University Children’s Hospital Zurich, Switzerland. These children were able to walk or use a manual wheelchair for household distances. Furthermore, they were between 4 and 20 years old, had cognitive abilities to follow instructions, had no wounds or other medical conditions that prevented sensor placement, and provided informed consent to participate in the study. The local ethics committee approved this study (BASEC Nr.: 2019-00487).

### Procedure & equipment

Participants were equipped with five ZurichMOVE sensor modules, containing a 3-axis accelerometer, a 3-axis gyroscope, and an altimeter [11]. The sensors were placed on both wrists, the sternum, and the thigh and ankle of the less-affected side with corresponding hook-and-loop straps (Figure 1). Additional sensors were placed on walking aids and the spokes of the wheelchair if applicable. Time synchronization between the sensors was achieved by a master-slave configuration using Bluetooth Low Energy.

**Figure 1.**
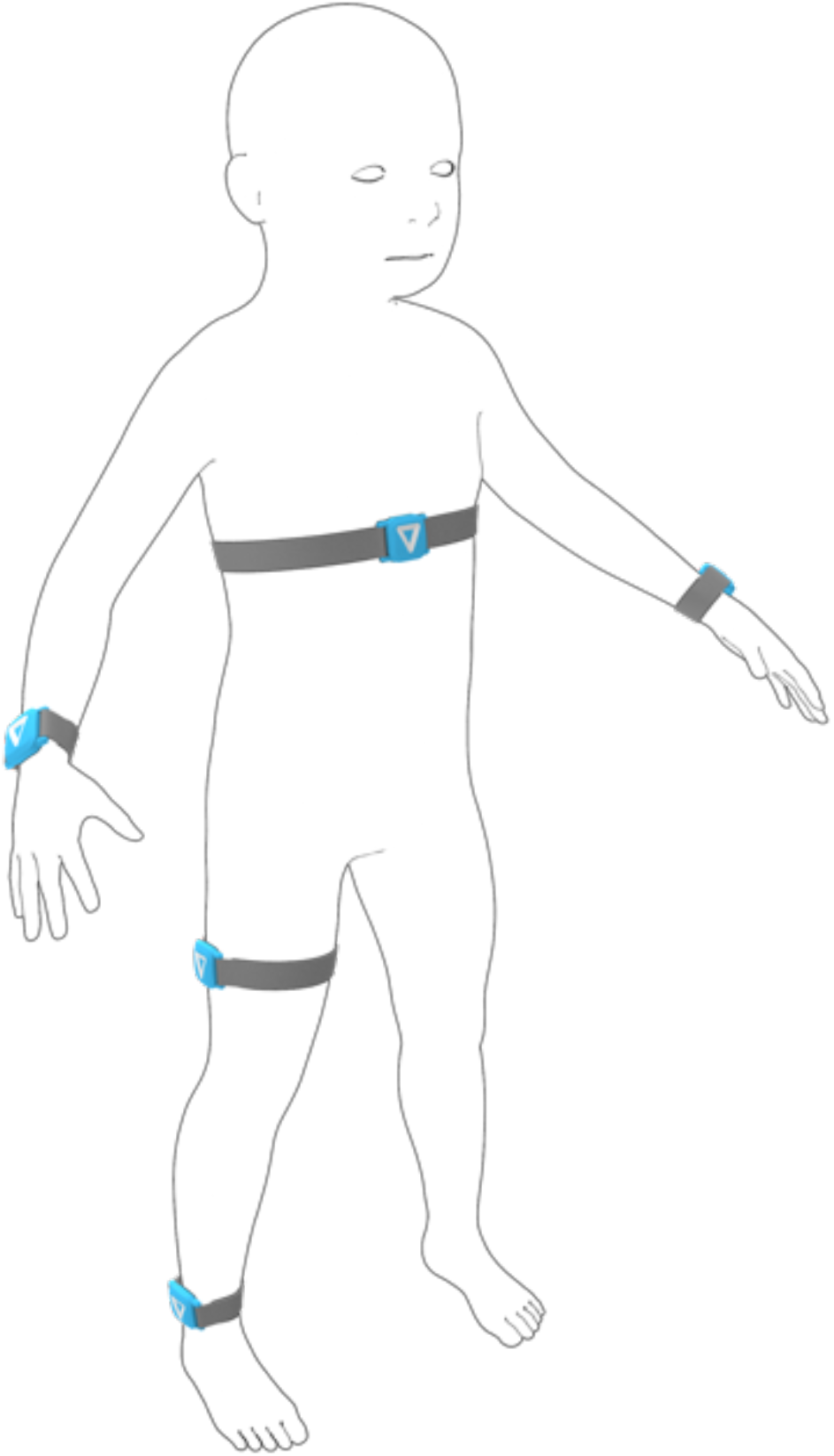
Placement of the five body-worn sensors at both wrists, the sternum, and the thigh and ankle of the less-affected side.

All participants performed a semi-structured activity circuit at the rehabilitation center. They watched a movie on a tablet in their bedroom, played a self-selected game (e.g., card games, puzzles, etc.) in the living room, drank a glass of water in the restaurant, cycled in the gym hall, and played what they wanted to on the outdoor playground (e.g., catching and throwing balls, swinging, etc.). Participants were encouraged to walk, wheel, climb stairs, and take the elevator between these facilities, depending on their functional abilities. No instructions were given on how to do these activities so that the children moved as they would in real life. To increase comparability with everyday life, the circuit covered activities of the target population that could be recognized by the algorithm and such that could not, to challenge its performance.

Video recordings from an external perspective served as reference criteria to determine the algorithm’s performance. The sampling rates of all devices were set to 50 Hz, and timestamps were synchronized with the children clapping their hands in front of the camera.

### Data processing

The three sub-algorithms are comprehensively described in supplementary file 1 and briefly summarized here:

1. The **posture detection algorithm** identifies lying, sitting, and standing positions with the orientation of the trunk and thigh sensors. It is assumed that both sensors are in a vertical orientation during standing and in a horizontal orientation during lying. In a sitting position, the thigh is usually oriented horizontally while the trunk remains vertical. The cut-point between the sensor’s horizontal and vertical orientation were trained with the current dataset and a leave-one-subject-out approach. This approach reflects the algorithm’s performance when applying it to a new subject without having training data of that subject as it would be the case in upcoming studies. **Outcome measures:** the algorithm derives the time spent in each position and the number of sit-to-stand transitions.
2. The **wheeling detection algorithm** discriminates between wheeling and non-wheeling periods with data of the wheelchair sensor and distinguishes between active and passive wheeling with the wrist sensor of the less-affected hand. The algorithm applies several thresholds to the angular rate of the wheel to detect wheeling periods [12]. Then, the wheeling periods are segmented into 5.12 s windows and an overlap of 75%. And finally, the orientation of the less-affected hand is used to classify active and passive wheeling. The cut-point was again trained with the current dataset and a leave-one-subject-out approach. **Outcome measures:** the algorithm derives the total duration of active and passive wheeling separately.
3. The **walking detection algorithm** uses walking-specific characteristics of the ankle’s gyroscope signal to discriminate between walking and non-walking periods. The acceleration signal of the sensor on the walking aid is used to distinguish between free and assisted walking. Moreover, the algorithm detects stair climbing periods with the altimeter of the ankle sensor. Whenever a child walks four consecutive steps and covers an altitude change between 7 and 49 cm per step, this period is considered stair climbing. Positive altitude changes are classified as ascending and negative ones as descending. **Outcome measures:** the algorithm derives the free and assisted walking duration and estimates the covered altitude change during stair climbing periods.

The sub-algorithms 1 & 3 were applied to data from all participants, while the wheeling detection algorithm was only applied to data from participants using a wheelchair.

Two researchers labeled the video recordings independently as lying, sitting, standing, and unknown (not visible). Sitting was sub-labeled as sitting, kneeling, being carried, active wheeling, passive wheeling, cycling, swinging, and sliding. Standing was sub-labeled as standing, free walking, assisted walking, going upstairs, going downstairs, and jumping.

Disagreements lasting more than one second were discussed retrospectively. In the case of consensus, the labels were corrected. Otherwise, the labels were retained. We published the labeled dataset and added a detailed definition of the individual activities [13].

### Statistical analysis

The algorithm’s activity detection accuracy was calculated by the proportion of correctly predicted data samples over all predictions. We further calculated the sensitivity and precision for each activity separately:

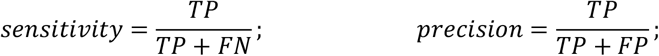

with TP = true positive predictions, FN = false negative predictions, and FP = false positive predictions. Unknown periods (0.4%), as well as periods with disagreement in the video labels (4.2%) and periods in which the sensors were not placed correctly (1.9%), were ignored in this analysis. An accuracy, sensitivity, and precision of >90% was considered excellent, 80% to 90% good, 70% to 80% moderate, and less than 70% weak [10].

Moreover, the agreement of the algorithm’s outcome measures with those of the reference system was determined with three different metrics. First, the measurement error was estimated as the difference between the algorithm’s measures and those derived from the video labels (reference values). The smaller difference to one of the two reference values was used. The measurement error was determined for each participant separately. The measurement error represents agreement within the activity circuit. Second, the relative measurement error was determined by dividing the measurement error by the mean of the reference values. Relative measurement error reflects the agreement of long-term measurements. Third, the Spearman’s rank correlation coefficient was calculated. These coefficients demonstrate the algorithm’s ability to discriminate between participants despite measurement error.

## Results

Twelve girls and 19 boys (11.8 ± 3.2 years) with various medical diagnoses and mobility impairments completed the study protocol. Their level of gross motor function, diagnosis, and use of mobility aids is shown in Figure 2A. The activity circuit lasted 46 min on average, and the performed activities depended on the participants’ capabilities. An overview of the dataset is illustrated in Figure 2B, and the resulting sample sizes of each activity are in line with previous studies [7–10]. Nineteen participants were able to walk with or without walking aids, eleven were wheelchair-dependent, and one participant moved around on a bicycle. Seven of those who were able to walk also used a wheelchair for longer distances.

**Figure 2.**
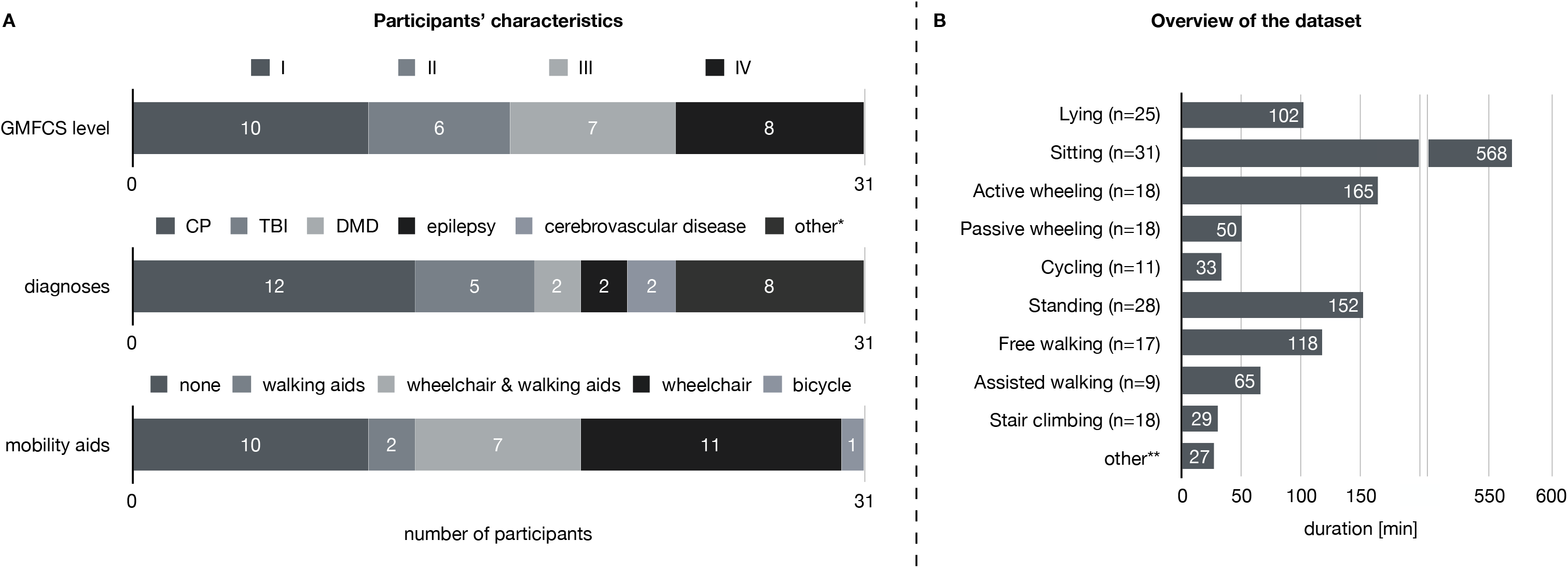
Illustration of the participants’ characteristics (A) and the collected dataset (B). GMFCS = gross motor function classification system; CP = cerebral palsy; TBI = traumatic brain injury; DMD = dissociative movement disorder; n = number of participants performing that activity. *chromosomal abnormality, congenital malformation syndrome, demyelinating disease, hereditary ataxia, multiple sclerosis, neoplasm, osteomyelitis & spina bifida. **kneeling (n=5, 13 min), being carried (n=1, <1 min), sliding (n=1, <1 min), swinging (n=4, 13 min) & jumping (n=2, 1 min).

The between-researcher agreement of the video labels was 96%. The majority of disagreement occurred due to uncertainty about discriminating lying and sitting when participants were seated in the bed with a backward-tilted backrest and discriminating standing and walking in participants making small and discontinuous steps.

The accuracy, sensitivity, and precision of the three sub-algorithms are presented in three corresponding confusion matrices in Figure 3. The posture detection algorithm revealed excellent performance. Sensitivities and precisions to detect lying, sitting, and standing were greater than 93%. In case of misclassification, the three postures were confused with each other but not with other activities, except for cycling. Roughly one-third of the cycling time was misclassified as standing. The wheeling detection algorithm revealed good to excellent performance, while the classification of active wheeling was more sensitive and precise than the classification of passive wheeling. Wheeling was confused with sitting and assisted walking but not with other activities. The walking detection per se revealed a sensitivity and precision of almost 90%, and the remaining 10% were mainly confused with standing. However, the discrimination between level walking and stair climbing was erroneous, resulting in a weak performance to detect stair climbing and decreased sensitivity and precision to detect level walking periods. Still, the distinction between free and assisted walking as well as between going up- and downstairs was almost perfect.

**Figure 3.**
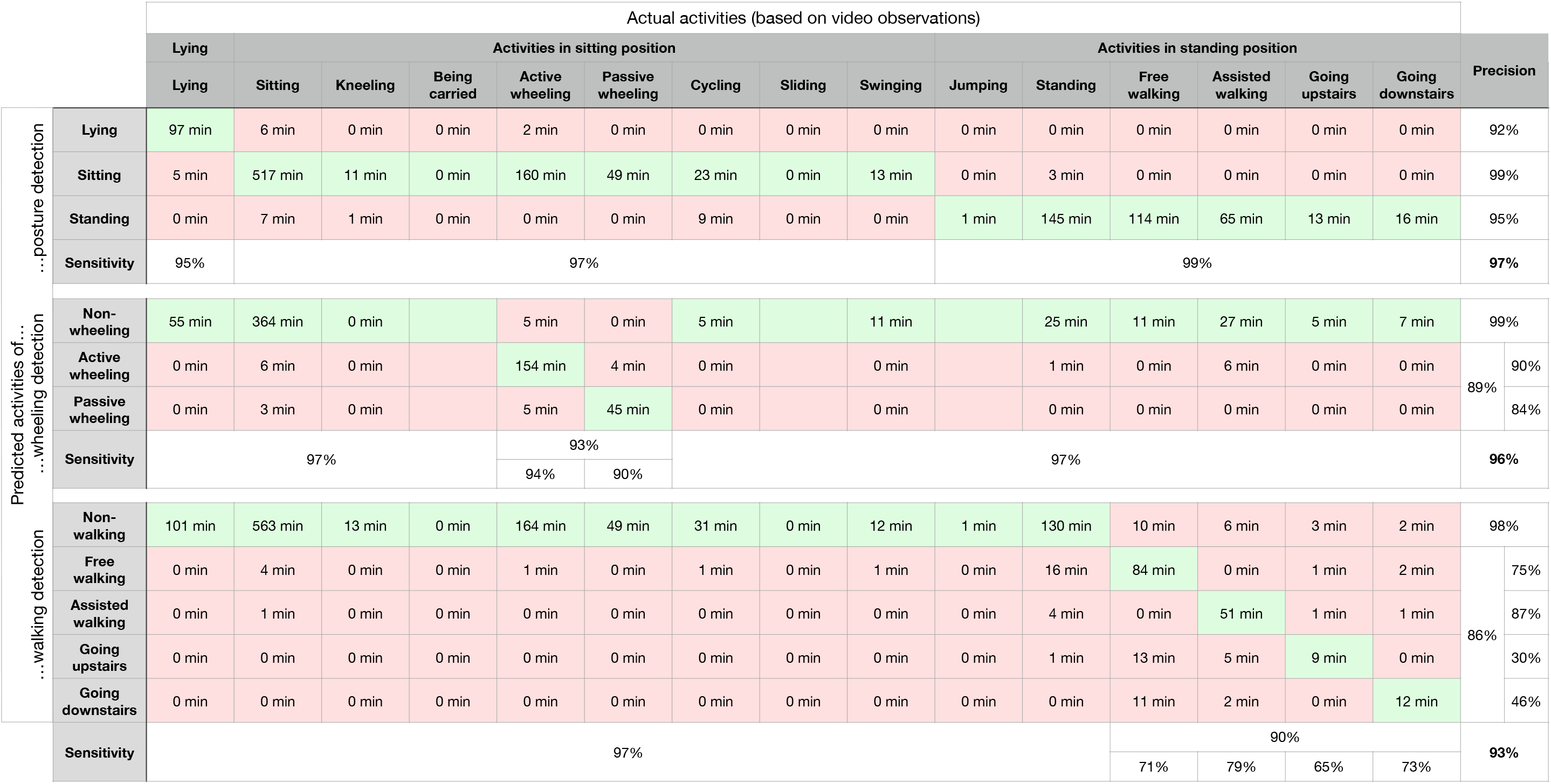
Confusion matrices of the three sub-algorithms.

The measurement errors of the outcome measures are depicted in Table 1. The algorithm estimated the duration of lying, sitting, standing, active wheeling, and walking with an error of less than 10% (interquartile range of relative difference between the algorithm’s measures and the reference values). The remaining performance measures revealed larger measurement errors. Systematic differences (median relative difference between the algorithm’s measures and the reference values) were smaller than 10%, except for standing up and stair climbing measures, which were systematically underestimated by the algorithm. The correlation coefficients ranged between .77 for the number of sit-to-stand transitions and .99 for the duration of sitting and standing periods.

**Table 1.** Measurement errors of the outcome measures

| *outcome measures* | *reference values* |  | *measurement error* |  | *relative measurement error* |  | *Spearman correlation* |
| --- | --- | --- | --- | --- | --- | --- | --- |
| | median | IQR | median | IQR | median | IQR | $\rho$ |
| ***duration of lying periods*** | 4.4 min | 1.7 min | 0.0 min | 0.1 min | 0% | 3% | 0.93 |
| ***duration of sitting periods*** | 27.8 min | 25.2 min | -0.1 min | 0.8 min | 0% | 5% | 0.98 |
| ***duration of standing periods*** | 14.0 min | 20.1 min | 0.0 min | 0.8 min | 0% | 6% | 0.99 |
| ***number of sit-to-stand transitions*** | 5 | 3 | -1 | 2 | -20% | 32% | 0.77 |
| ***duration of active wheeling*** | 9.9 min | 4.8 min | 0.0 min | 0.5 min | 0% | 5% | 0.91 |
| ***duration of passive wheeling*** | 2.1 min | 3.0 min | 0.0 min | 0.3 min | 2% | 19% | 0.84 |
| ***duration of all walking periods*** | 10.7 min | 5.2 min | 0.1 min | 0.9 min | 0% | 8% | 0.94 |
| *duration of free walking periods* | 8.7 min | 9.8 min | 0.0 min | 1.5 min | -3% | 41% | 0.91 |
| *duration of assisted walking periods* | 7.0 min | 9.0 min | 0.2 min | 2.2 min | 4% | 26% | 0.93 |
| *altitude change during going upstairs\** | 6.6 m | 8.2 m | -0.7 m | 2.4 m | -23% | 52% | 0.89 |
| *altitude change during going downstairs\** | 6.6 m | 8.2 m | -1.0 m | 0.8 m | -27% | 32% | 0.97 |

\*this analysis was limited to indoor activities since it was impossible to determine the altitude change during slope walking periods occurring only outdoors. Indoors, the heights of each flight of stairs were known and the covered flights of stairs were counted in the video recordings.

## Discussion

This study introduces and validates a novel algorithm to derive clinically relevant performance measures based on wearable inertial sensor data of children with mobility impairments. The algorithm performed excellent in detecting lying, sitting, and standing; good to excellent in detecting general walking periods as well as active and passive wheeling periods; and weak in discriminating between level walking and stair climbing.

The algorithm confused lying with sitting mainly when the children were lying while resting on their elbows and thus, having a relatively upright trunk orientation. Conversely, false positive lying detections occurred predominantly when the children were sitting and leaning forward (e.g., to pick up an object from the floor or lock the wheels of the wheelchair). These confusions can easily be explained since the algorithm relies on the orientation of the trunk to classify lying and sitting positions. Furthermore, we expect that these confusions hardly affect the overall lying and sitting duration of long-term measurements.

Cycling was often classified as standing rather than sitting, which can also be explained by the algorithm using the orientation of the thigh to discriminate between sitting and standing positions. In children with a lot of daily cycling activities, this would lead to overestimating weight-bearing activities, especially in wheelchair-dependent children. We suggest developing an algorithm that detects cycling periods specifically or using a protocol reporting daily cycling activities to overcome this limitation.

During the circuit, it happened that the trunk and thigh sensors slipped downward, and we replaced the sensors as soon as we realized it. These periods were detected in the video recordings and ignored in the data analysis since we intended to validate the algorithm and not the slip resistance of the straps. However, this can also happen in daily life and would lead to erroneous data. Therefore, sensors should be placed solidly on the children, and the families should be instructed to verify the sensor positions regularly.

If at all, sitting and assisted walking were misclassified as wheeling and decreased the precision of the wheeling detection algorithm. Wheeling periods in the video recordings were identified independent of the covered distance and underlying velocity, since this cannot be observed accurately. In contrast, the algorithm only detected wheeling periods in which the wheel exceeded an angular rate of 10°/s and a turn of 80° [12]. This discrepancy could explain the confusion between sitting and wheeling. The confusion between assisted walking and wheeling occurred in a single participant. He was walking while we pushed his wheelchair alongside. This can happen in daily life as well, especially during transitions between wheeling and walking periods. However, we believe that these periods can be neglected compared to the duration of wheeling and walking in long-term measurements.

Periods of going up- and downstairs were detected with a weak precision of 30% and 46%. Free walking was often confused with stair climbing. In the reference system, we did not discriminate between level and slope walking, even though some children walked outdoors on slopes corresponding to three flights of stairs. Reducing the analysis to indoor periods revealed a higher precision of 44% and 60%. Still, stair climbing was often confused with walking and standing. More severely impaired children took adjusting steps when ascending and descending stairs and did small breaks on each step. The algorithm failed to detect these stair climbing patterns, and four consecutive steps are required to detect stair climbing with the current algorithm. Despite classification inaccuracy, the algorithm can still discriminate between participants with low and high stair climbing activity, indicated by correlation coefficients of 0.89 and 0.97.

A comparison of the results with previous literature is difficult due to the heterogeneity in study designs. There are dissimilarities in the study population, the measurement devices, the performed activity protocols, and the number and type of detected activities by the algorithms. Still, our novel algorithm outperforms previous algorithms validated in children with mobility impairments [7–10]. This is remarkable for three main reasons. First, our study population has a larger variety in medical diagnoses and levels of impairments, which challenges the algorithm to find a solution that fits all. It has been shown that subgroup-specific algorithms or fully-personalized approaches reveal higher accuracies than those covering the whole population [14]. Second, none of the studies mentioned above included stair climbing as an activity of interest, and excluding stair climbing from our analysis would further increase the accuracy of our algorithm since stair climbing detection revealed the least accurate results in our study. And third, we determined the algorithm’s performance with the whole dataset, while the other studies excluded or ignored transitions between activities [7–9] or disregarded activities lasting less than five seconds [10]. We argue that transitions and short-lasting activities are challenging to detect, and they should be included in the analysis to reflect real-life data to increase the external validity of the study results.

Moreover, our study protocol included activities not classified by the algorithm to challenge activity detection and reflect everyday life activities, which is another strength of our study protocol. However, it has to be shown whether the amount of performed activities in our dataset reflects daily activities of children with mobility impairments, and it remains questionable if the results of this study can be transferred to long-term measurements in daily life. Hence, we encourage the research community to develop methodologies to validate wearable sensor technology and their underlying algorithms in the children’s real world and not just during semi-structured activity circuits, even though the latter is recommended as a standard for such validity studies [15].

## Conclusion

This study introduces three sub-algorithms that determine clinically meaningful outcome measures based on wearable inertial sensor data in the daily life of children with mobility impairments. The first algorithm determines the duration of lying, sitting, and standing as well as the number of sit-to-stand transitions with two sensors placed on the trunk and the thigh. The second algorithm measures the duration of active and passive wheeling periods with a wrist sensor and a sensor placed on the spokes of the wheelchair. And the third algorithm determines the duration of free and assisted walking as well as the altitude change covered during stair climbing periods with an ankle sensor and a sensor placed on walking aids. The sub-algorithms are well described, can be reproduced, and applied to other inertial sensor technologies. Moreover, they were validated in children with mobility impairments and can be used in clinical practice and clinical trials to determine the children’s motor performance in their habitual environment. Besides, we published the labeled dataset enabling the evaluation of future algorithms.

## Data Availability

All data produced in the present study are available upon reasonable request to the authors.

## Acknowledgements

We would like to thank all participants for their participation.

## Notes

**Funding:** This study was supported by the Walter Muggli Fund of the ACCENTUS Foundation, Zurich, Switzerland, the Children’s Research Center of the University Children’s Hospital of Zurich, Switzerland, and the Anna Mueller Grocholski Foundation, Zurich, Switzerland. The funding sources were not involved in designing the study; in collecting, analyzing, and interpreting data; and in writing the report.

### Competing Interest Statement

The authors have declared no competing interest.

### Funding Statement

This study was supported by the Walter Muggli Fund of the ACCENTUS Foundation, Zurich, Switzerland, the Children's Research Center of the University Children's Hospital of Zurich, Switzerland, and the Anna Mueller Grocholski Foundation, Zurich, Switzerland. The funding sources were not involved in designing the study; in collecting, analyzing, and interpreting data; and in writing the report.

### Author Declarations

The ethics committee of the Canton Zurich, Switzerland (Kantonale Ethikkommission Zuerich) gave ethical approval for this work.

