## supplementary file 1 for "Accuracy of sensor-based classification of clinically relevant motor activities in daily life of children with mobility impairments"

### Supplementary file 1 - posture and mobility detection algorithm

Fabian Marcel Rast[a,b,c]

[a] Swiss Children's Rehab, University Children's Hospital Zurich, Mühlebergstrasse 104, 8910, Affoltern am Albis, Switzerland
[b] Children's Research Center, University Children's Hospital of Zurich, University of Zurich, Zurich, Switzerland
[c] Rehabilitation Engineering Laboratory, Department of Health Sciences and Technology, ETH Zurich, Zurich, Switzerland

**Abstract**

The algorithm can be divided into three independent parts using different sensor setups:

1. The posture detection algorithm detects lying, sitting, and standing positions based on data of the trunk and thigh sensors.
2. The wheeling detection algorithm detects wheeling periods with data of the wheelchair sensor and discriminates between active and passive wheeling with data of the wrist sensor of the dominant hand.
3. The walking detection algorithm detects walking periods and differentiates between level walking and stair climbing with data of a single ankle sensor. Further, the algorithm discriminates between free and assisted walking with data of the sensor attached to a walking aid.

#### 1. Raw data

We used the data of inertial measurement units containing a 3-axis accelerometer, a 3-axis gyroscope, and a barometric pressure sensor, as well as Bluetooth Low Energy for time synchronization (see Figure 1).[1] However, the algorithm can be applied to any measurement unit containing the required sensor modalities. The raw data needs to be resampled to 50 $Hz$, and the signals must be measured in or converted to the following units:

- acceleration $a \Rightarrow m/s^2$
- angular rate $\omega \Rightarrow \ ^{\circ}/s$
- barometric pressure $p \Rightarrow Pa$

#### 2. Posture detection algorithm

This part of the algorithm detects lying, sitting, and standing positions based on data of the trunk and thigh sensors.

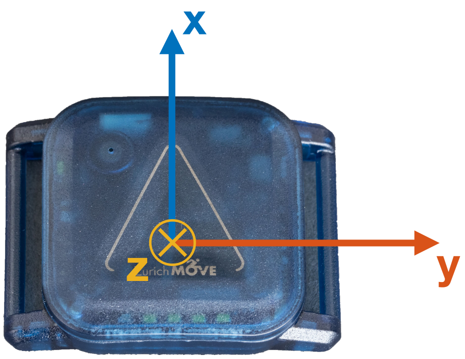


Figure 1: The ZurichMOVE sensor and its coordiante system (created by Rehabilitation Engineering Laboratory, ETH Zurich).

##### 2.1. Sensor placement

The trunk sensor needs to be placed on the sternum with the x-axis facing toward the belly button. The thigh sensor needs to be placed mid-thigh on the lateral side of the less-affected leg. Here, the x-axis faces toward the knee (see Figure 2).

##### 2.2. Orientation estimation

Before estimating the orientation of the sensor, the algorithm corrects the offset and drift of the gyroscope signal.[2] First, still phases are detected by applying a 2nd order high-pass filter (cut-off frequency $= 0.5\ Hz$), a low-pass filter (cut-off frequency $= 2\ Hz$), and a threshold of $1\ ^{\circ}\backslash s$.[3] Then, the drift of the gyroscope signal is estimated by piecewise low-pass filtering of each axis, linearly interpolating between the still phases, and limiting the slew rate of the signal to $500\ \mu^{\circ}\backslash s\backslash s$. And finally, this drift is subtracted from the raw gyroscope measurements.

To estimate the orientation of the sensor, the acceleration and the corrected gyroscope signals are fused with the open-source algorithm of S. Madgwick.[4] The filter gain $\beta$ was set to 0.03 which provided optimal performance in previous experiments.[4]. The output is a vector containing the quaternions of each sample $\vec{q} = [q_0\ q_1\ q_2\ q_3]^T$. In the neutral position ($\vec{q} = [1\ 0\ 0\ 0]^T$), the z-axis points towards the floor.

As the last step, the pitch angle of the sensor's orientation is derived from the quaternions. The pitch angle is defined as the deviation of the sensor's orientation from its neutral position around a new y-axis after rotating the sensor around its z-axis. It is calculated with the following equation:

$$\varphi = \arcsin\Big(2(q_0 q_2 - q_3 q_1)\Big) * \frac{180}{\pi}$$

This angle is filtered using a 5th order low-pass filter (cut-off frequency $= 0.1\ Hz$). An angle of $0^{\circ}$ represents a horizontal orientation, while an angle of $\pm 90^{\circ}$ represents a vertical orientation. Negative values result when the x-axis of the sensor points downward, and positive values result when the x-axis points upward. The former corresponds to a standing position, while the latter would correspond to a handstand position. Signals of which the mean of the whole measurement period exceeds $0^{\circ}$ are multiplied by -1 since we assume that the sensor was placed upside down rather than

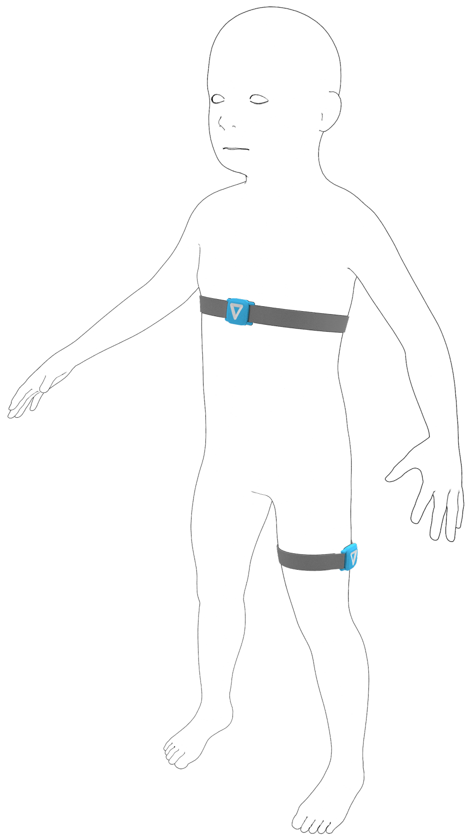

Figure 2: Sensor placement of the posture detection algorithm (created by Rehabilitation Engineering Laboratory, ETH Zurich).

the participant being in this position for a prolonged time.

##### 2.3. Classification of lying, sitting, and standing

In lying, both sensors are horizontal while they are vertical during standing. In sitting, however, the thigh sensor is horizontal and the trunk sensor is vertical. A vertical thigh sensor and a horizontal trunk sensor is uncommon and probably reflects a standing position with bending forward. Hence, the algorithm classifies this scenario as standing. The thresholds to distinguish between a horizontal and vertical orientation were trained with labeled data of children with mobility impairments and a decision tree by minimizing the Gini's Diversity Index. The resulting thresholds are $T_{trunk} = -35.9°$ and $T_{thigh} = -48.4°$.

##### 2.4. Outcome measures

After detecting lying, sitting, and standing positions, the algorithm determines the duration the participant spent in each position throughout the measurement period. Moreover, the number of transitions between a sitting and a standing position are counted. The minimal duration between two consecutive sit-to-stand transitions was set to 2 *min* to avoid an overestimation in noisy data or during cycling periods.

#### 3. Wheeling detection algorithm

This part of the algorithm detects wheeling periods with data of the wheelchair sensor and discriminates between active and passive wheeling with data of the wrist sensor of the dominant hand.

##### *3.1. Sensor placement*

The wheelchair sensor needs to be placed on the spokes of the wheelchair, with the z-axis being parallel to the axis of the wheel. The direction of the z-axis does not matter since the algorithm assumes that the participant more frequently wheels forward than backward. The wrist sensor is worn on the dominant hand as a watch. The x-axis faces toward the fingers (see Figure 3). The user selects the dominant hand by placing a single sensor on the corresponding wrist. If data of both wrist sensors are available, the algorithm uses the side which reveals a higher acceleration magnitude during wheeling periods.

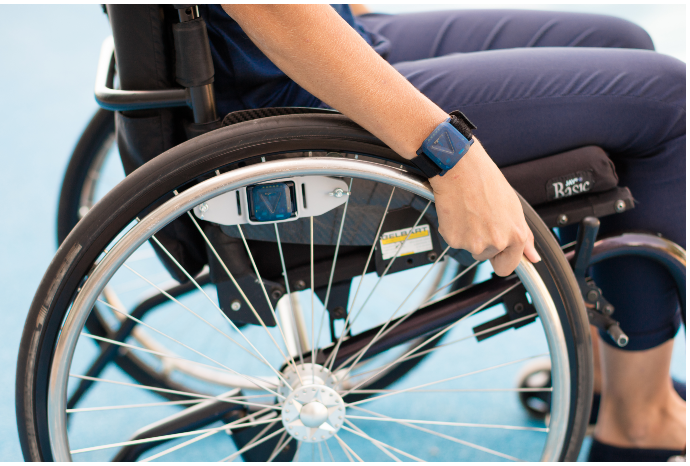

Figure 3: Sensor placement of the wheeling detection algorithm (created by Rehabilitation Engineering Laboratory, ETH Zurich).

##### *3.2. Classification of non-wheeling activities, active wheeling and passive wheeling*

This part of the algorithm is an adapted version of a previously published algorithm that was developed for patients with a spinal cord injury.[5] The feature selection process was repeated with data of children with mobility impairments and the resulting algorithm is described in the following sections.

*3.2.1. Detection of wheeling periods*

This part depends solely on the z-axis of the wheelchair sensor $a_{wheel,z}$ and $\omega_{wheel,z}$. As a first step, it is verified if the z-axis of the sensor is in a horizontal orientation which is the case if the sensor is fixed to the spokes of the wheel. In this case, the acceleration signal due to gravity is close to zero. In contrast, the signal is close to 9.81 $m/s^2$ if the sensor is lying around in neutral position. Therefore, periods in which $a_{wheel,z}$ is $> 0.5 * 9.81$ $m/s^2$ for longer than 1 $min$ are classified as non-wheeling activities and ignored during the following steps. Before applying this cut-off, the signal is processed with a low-pass filter (cut-off frequency = 0.05 $Hz$).

Then, plateaus in the gyroscope signal of five samples in a row or longer are set to zero to remove gyroscope data with low quality. The resulting signal is used to detect wheeling periods in three steps: identifying preliminary wheeling periods, classifying them as valid and invalid wheeling periods, and fusing valid wheeling periods by analyzing the rest phases between two consecutive wheeling periods. First, a threshold ($|\omega_{wheel,z}| > 0.4°/s$) is applied to identify preliminary wheeling periods. Second, for each period, the following heuristic rules are used to detect valid wheeling periods:

- $\max |\omega_{wheel,z}| > 10°/s$
- $Var(\omega_{wheel,z}) > 1°/s$
- $\int |\omega_{wheel,z}| dt > 80°$

Third, valid wheeling periods that are less than 2 $s$ apart are fused. In addition, rest phases between two valid wheeling periods that contain more than 80% of preliminary wheeling periods as well as those that are shorter than 0.8 $s$ are also classified as a valid wheeling period. Finally, valid wheeling periods that are less than 2 $s$ apart are fused again.

*3.2.2. Discrimination between active and passive wheeling*

As a first step, the raw data of the wrist sensor is filtered. The acceleration signal is passed through an infinite impulse response eight order elliptic low-pass filter with a cut-off frequency of 0.3 $Hz$, a passband ripple of 0.02 $dB$, and a minimum stopband attenuation of 200 $dB$ in order to separate the static acceleration component due to gravity $a_{static}$ from the dynamic acceleration component resulting from wrist movement $a_{dynamic}$.[6]

Wheeling periods lasting longer than 5.12 $s$ are divided into segments with a window length of 5.12 $s$ and an overlap of 75%. Each segment in which the wrist sensor is not able to communicate with the wheelchair sensor via Bluetooth Low Energy is classified as non-wheeling activity. Here, it is assumed that the wheelchair is far away from the participant. The remaining segments are either classified as active or passive wheeling. The same features of the original publication of this algorithm [5] were calculated and the feature selection process was repeated with data of children with mobility impairments. It revealed just a single relevant feature: $P_{10th}(a_{wrist,static,x})$. This feature is a surrogate for the orientation of the wrist. It is 9.81 $m/s^2$ if the z-axis is parallel to gravity and zero if the z-axis is perpendicular to gravity. Movement related features did not improve classification accuracy since we encouraged children to do hand activities while they were pushed around in their wheelchair. The threshold to distinguish between active and passive wheeling was trained with a decision tree by minimizing the Gini's Diversity Index. The resulting threshold is $T_{wrist} = -0.61 * 9.81$ $m/s^2$. Wheeling periods with the hand facing down towards the wheel are classified as active wheeling while periods with the hand facing more horizontal or upward are classified as passive wheeling.

##### *3.3. Outcome measures*

The algorithm derives the total duration of active and passive wheeling separately.

#### 4. Walking detection algorithm

This part of the algorithm detects walking periods and differentiates between level walking and stair climbing with data of a single ankle sensor. Further, the algorithm discriminates between free and assisted walking with data of the sensor attached to a walking aid.

##### *4.1. Sensor placement*

The ankle sensor is worn on the less-affected ankle. The x-axis faces toward the floor and points to the lateral malleolus (see Figure 4). If applicable, a sensor is placed firmly on the walking aid of the participant. The position and orientation of this sensor are irrelevant.

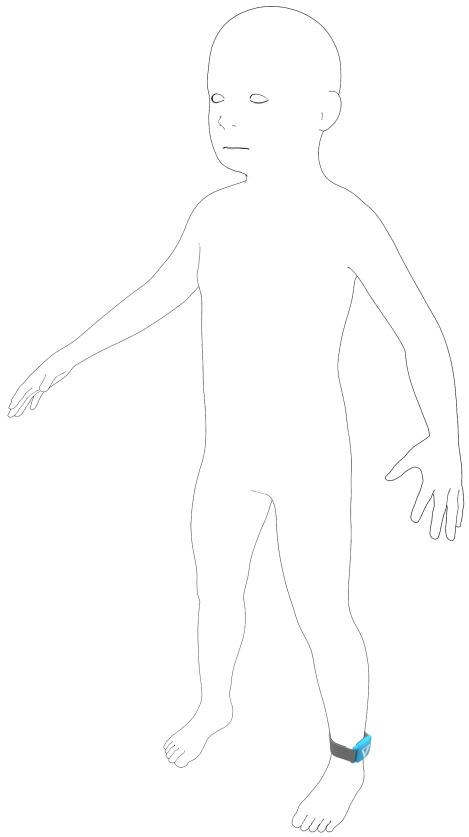

Figure 4: Sensor placement of the walking detection algorithm (created by Rehabilitation Engineering Laboratory, ETH Zurich).

##### 4.2. Detection of walking bouts

###### 4.2.1. Preprocessing I

As a first step, the algorithm verifies the placement of the ankle sensor. If the average acceleration signal of the x-axis is greater than zero, it is assumed that the sensor was placed upside down. In this case, the sensor is rotated 180° around its z-axis by multiplying the acceleration and gyroscope signals of the x- and y-axis by $-1$. Then, the bias and drift of the gyroscope signal are corrected as described in chapter 2.2

###### 4.2.2. Segmentation and preprocessing II

A 5th order low-pass filter (cut-off frequency $= 3\ Hz$) is applied to the gyroscope signal. Then, the signal is segmented into windows of 30 $s$ and an overlap of 15 $s$. In each segment, the angular rate around the mediolateral axis $\omega_{ml}$ is determined by correcting for misalignment around the x-axis of the ankle sensor and by the assumption that the majority of leg movement occurs in the sagittal plane:

$$\begin{pmatrix} \cdot \\ \omega_{ml} \end{pmatrix} = \mathbf{v} \begin{pmatrix} \omega_y \\ \omega_z \end{pmatrix},$$

with $\mathbf{v}$ being the eigenvector of $cov(\omega_y, \omega_z)$ with the largest eigenvalue. Since the eigenvector can point in both directions, the signal $\omega_{ml}$ has to be multiplied with $-1$ whenever it is upside down. To verify this, the algorithm uses the fact that the angular rate is larger during the swing phase compared to the stance phase. It compares the means of the upper and lower envelops of the signal and multiplies $\omega_{ml}$ with $-1$ whenever the mean of the lower envelop is larger than the the mean of the upper envelop. Consequently, positive values of $\omega_{ml}$ correspond to a backward rotation of the shank as during the swing phase and vice versa. An exemplary signal is shown in Figure 5

###### 4.2.3. Step detection

The algorithm detects steps by finding local maxima in $\omega_{ml}$, corresponding to the peak angular rate during mid-swing of each step (see Figure 5). The amplitude of these maxima, as well as the duration between two consecutive maxima, must exceed the thresholds $T_{peakheight}$ and $T_{peakdistance}$, respectively. These thresholds are adapted to the underlying data of each segment and, thus, to individual gait patterns.

$$T_{peakheight} = max(50°/s, 0.2P_{99th}(\omega_{ml}))$$

A minimum of $50°/s$ was chosen to exclude maxima of non-walking data.[7]

$$T_{peakdistance} = \frac{0.5}{\tilde{f}_{walking}}$$

with $\tilde{f}_{walking}$ being the median estimated step frequency (steps per second) of each segment. An initial step frequency $f_{initial}$ is estimated for the whole segment by applying a fast Fourier transformation to $\omega_{ml}$ and taking the first main frequency component. An adapted step frequency is estimated by repeating this step in a sliding window of $\frac{3}{f_{initial}}$ and an overlap of $\frac{2}{f_{initial}}$. Eventually, a moving average filter with a span of $\frac{9}{f_{initial}}$ is applied to determine the time-dependent step frequency $f_{walking}$. Eventually, only steps of the middle 15 $s$ of each segment are considered to avoid duplicates in overlapping segments.

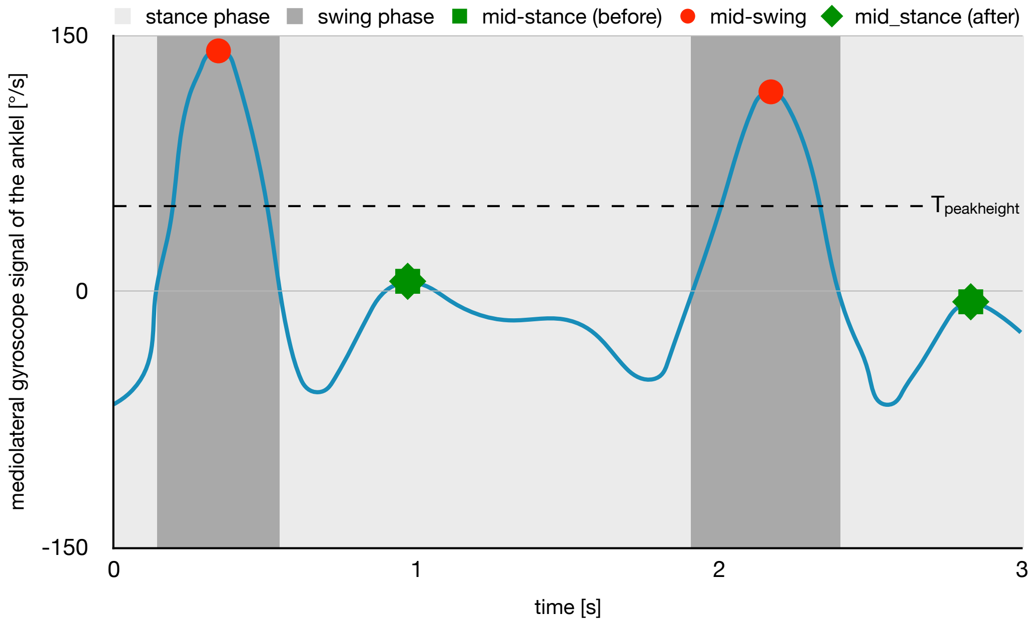


Figure 5: Exemplary illustration of the gyroscope signal of two steps as well as the corresponding gait phases and gait events detection.

**Removing unreasonable steps** The swing phase of each step is defined as the time $t_{swing}$ between the first zero-crossings of $\omega_{ml}$ before and after mid-swing. The stance phase is defined as the time $t_{stance}$ between the first zero-crossing of $\omega_{ml}$ after the preceding mid-swing and the beginning of the current swing phase. Steps with $t_{swing} < 100\ ms$ or $t_{stance} < 200\ ms$ are not considered valid steps.[8]

###### *4.2.4. Mid-stance detection and classification of walking periods*

The algorithm detects the mid-stance before and after each mid-swing detected above, and the period between the two mid-stance is classified as walking. During continuous gait, the mid-stance after one mid-swing is equal to the mid-stance before the subsequent mid-swing and the whole period is classified as walking. During interrupted gait, the two mid-stance do not overlap, and the period between is classified as non-walking (see Figure 6). Mid-stance is defined as the time of the largest local maximum in $\omega_{ml}$ during the stance phase (see Figure 5 and Figure 6). The occurrence of local maxima is a typical characteristic of gait. If there is no local maximum (e.g., during cycling periods), the algorithm removes the corresponding step. Moreover, the angular rate of the local maxima is usually negative which corresponds to a forward progression of the shank. However, the angular rate can reach positive values during walking on uneven surfaces or stair climbing. Still, the local maxima during the stance phases are considerably smaller than those during the swing phases. Therefore, the algorithm removes steps whenever the local maximum during the stance phase exceeds half of the maximum during the swing phase. Eventually, $t_{stance}$ needs to be smaller than $\frac{3}{\bar{f}_{walking}}$. Otherwise, the mid-stance after the preceding step is set to the end of the preceding swing phase, and the mid-stance before the subsequent step is set to the beginning of the subsequent step. Consequently, the stance phase will be classified as a non-walking period.

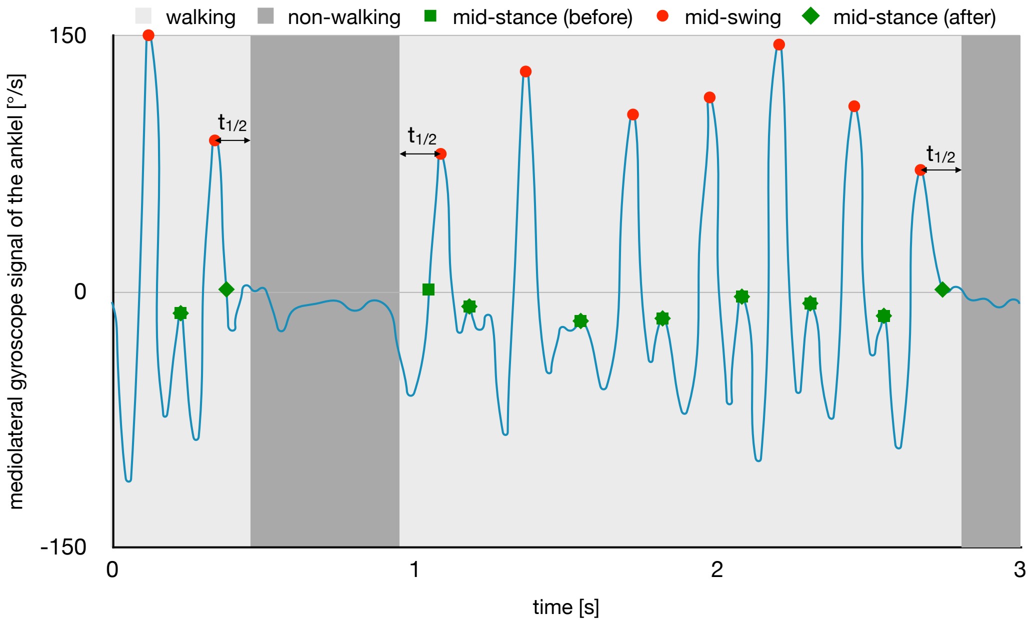


Figure 6: Exemplary illustration of the gyroscope signal of interrupted gait as well as the corresponding gait events detection and the resulting walking classification. $t_{1/2}$ represents half of the mean step duration of each walking period to determine the corresponding start and end points

**Removing unreasonable steps** The algorithm determines the orientation of the ankle sensor $\varphi_{ankle}$ (see chapter 2.2) between the mid-stance before and after each step. Then, steps with unreasonable orientation and range of motion are classified as non-walking periods if one of the following criteria is fulfilled:

$$\begin{array}{rcc} min(\varphi_{ankle}) & > & -45° \\ max(\varphi_{ankle}) & > & 0° \\ max(\varphi_{ankle}) - min(\varphi_{ankle}) & < & 5° \\ max(\varphi_{ankle}) - min(\varphi_{ankle}) & > & 90° \end{array}$$

##### *4.2.5. Break detection*

This part of the algorithm detects breaks within each walking period and classifies them as non-walking. It is assumed that the step duration between two consecutive mid-swings remains relatively constant during continuous gait. Therefore, breaks are detected with long and irregular step durations. The specific criteria depend on the number of steps within each walking period and are defined as follows:

**≥4 steps** First, the algorithm calculates the median step duration of four consecutive steps. If the step duration of one of these steps is greater than one and a half times the median step duration, the corresponding stance phase is classified as non-walking. This part is repeated for each set of four consecutive steps.

**3 steps** If one of the two step durations is more than twice as long as the other, the whole period is classified as non-walking.

**2 steps** If the step duration is longer than 5 $s$, the whole period is classified as non-walking.

**1 step** Walking periods with a single step are ignored and classified as non-walking.

*4.2.6. Start and end point*

At the beginning and end of each walking period, there is no typical mid-stance. Hence, each walking period begins half of the mean step duration before the mid-swing of the first step and ends at half of the mean step duration $t_{1/2}$ after the mid-swing of the last step (see Figure 6).

*4.3. Use of walking aids*

The algorithm classifies each walking period as either free walking or assisted walking. If the participant does not use a walking aid and there is no data available, all walking periods are classified as free walking. Walking periods in which the ankle sensor is not able to communicate with the sensor on the aid via Bluetooth Low Energy are classified as free walking, too. Here, it is assumed that the walking aid is far away from the participant. To determine whether the walking aid was used or not, the acceleration signal of the sensor placed on the walking aid is processed with a high-pass filter and a cut-off frequency of 0.3 $Hz$ to remove the gravity component in the signal. Then, for each walking period, the algorithm verifies if the 95th percentile of the magnitude of the filtered signal $a_{aid}$ is above a predefined threshold $T_{aid} = 0.05 * 9.81\ m/s^2$ to determine whether the walking aid was moved around or not:

$$P_{95th}\left(\sqrt{a_{aid,x}^2 + a_{aid,y}^2 + a_{aid,z}^2}\right) \begin{cases} \leq T_{aid} & \implies \text{free walking} \\ > T_{aid} & \implies \text{assisted walking} \end{cases}$$

*4.4. Detection of stair climbing*

The algorithm detects stair climbing periods based on the altitude change per step. Previously detected walking periods (independent of the use of walking aids) are classified as level walking, going upstairs, or going downstairs.

*4.4.1. Altitude estimation*

The pressure signal $p$ is transformed to the altitude above sea level $h$ with the following formula[1]:

$$h = \log \frac{1013}{p} * 7990$$

Then, a median filter with a window length of five samples is applied. The resulting signal is filtered with an 8th order decomposition, heuristic, automatic 1-D de-noising filter using a soft threshold and symlet8 wavelet.[9]

[1]a simplification of the international barometric formula

###### 4.4.2. Expected altitude change per step

to 42 *cm* since data of a single ankle sensor is used and participants can walk in a step-by-step or a step-over-step pattern. The algorithm adds a margin of 7 *cm*, which corresponds to the half of the smallest expected step height. Therefore, the lower border for discriminating between level walking and stair climbing was set to 7 $cm/step$, and the upper border was set to 49 $cm/step$. An upper border is needed as large altitude changes can occur when the environmental temperature changes rapidly (e.g., when walking out of a heated building).

###### 4.4.3. Classification of going upstairs and going downstairs

Walking periods containing less than four steps are always classified as level walking. The remaining walking periods are segmented into windows of four consecutive steps and an overlap of three steps. For each window, the algorithm determines the altitude change and compares it to the expected altitude change described above:

$$\begin{aligned} 7\ cm/step < \tfrac{\Delta h}{4\ steps} < 49\ cm/step \quad &\Longrightarrow \quad going\ upstairs \\ -49\ cm/step < \tfrac{\Delta h}{4\ steps} < -7\ cm/step \quad &\Longrightarrow \quad going\ downstairs \end{aligned}$$

##### 4.5. Outcome measures

Eventually, the algorithm derives the free and assisted walking duration and estimates the covered altitude change during stair climbing periods.

#### 5. Acknowledgments

This algorithm is an extension of previous algorithms developed at the Rehabilitation Engineering Laboratory, ETH Zurich (RELab). Therefore, I want to express my gratitude to three former and current members of the RELab for providing me with the codes of their work. Specifically, I want to thank Kaspar Leuenberger for providing his codes regarding the preprocessing of sensor data, Werner Popp for providing his wheeling detection algorithm for adults with a spinal cord injury, and Charlotte Werner for providing her codes regarding gait detection. Moreover, I want to thank Charlotte for revising this manuscript.
